# Using Bayesian Evidence Synthesis to estimate the number of sex workers in the United Kingdom

**DOI:** 10.64898/2026.05.21.26353767

**Authors:** Harry Long, Leonardo Gada, Lois Murray, Timothy Laurence, Andrew Hayward, Thomas Finnie

## Abstract

Sex work is diverse and includes a broad range of people and settings. Over the last thirty years, a large proportion of public health emergencies of international concern (PHEIC) have involved infections transmitted through sexual or close contact and in sexual networks (WHO 2024). Sex workers can face increased disadvantage in relation to these public health emergencies. Given the significant health inequalities sex workers can face, they should be eligible to receive targeted and tailored health support to reduce health protection risks (Hester 2019; Jeal and Salisbury 2004a). However, they are often not explicitly eligible for targeted and tailored support due to a lack of information on incidence, prevalence of disease, and even more basic data such as reliable estimates of the number of sex workers in the UK. Accordingly, the aim of this paper is to determine a population size estimate, with uncertainty, that is more robust than those currently available. In this study, we apply Bayesian Evidence Synthesis to bring together historic estimation efforts with recent ONS National Population Estimates and Genito-Urinary Medicine Clinics Attendance Data (GUMCAD) from the UK Health Security Agency (UKHSA). A key feature of our model is the embedding of uncertainty from each input study in model priors, hence propagating it through to our final estimate. The Bayesian evidence synthesis model estimated a total of 84,000 sex workers in the United Kingdom (95% credible interval: 49,000-130,000), representing 0.121% of the current UK population.

## 1 Introduction

Sex work is diverse and includes a broad range of people and settings. While any precise definition of sex work will inevitably be subject to counterexamples, an accepted characterisation is ‘the provision of sexual or erotic acts or sexual intimacy in exchange for payment or other benefit or need’ (Hester 2019, p.7). ‘Sex workers’ (hereinafter, ‘SWs’) is the term used for professionals engaging in consensual sex work. Sex workers can face significant health inequalities due to multiple challenges including barriers to accessing healthcare and experiences of stigma and discrimination (NUM 2023).

Over the last thirty years, a large proportion of public health emergencies of international concern (PHEIC) have involved infections transmitted through sexual or close contact and in sexual networks (WHO 2024). Sex workers can face increased disadvantage in relation to these public health emergencies, although it is important to recognise the wide diversity in sex work in relation to type and location. Not all people who are sex workers will have increased frequency of sexual contacts or be part of highly connected sexual networks, but those who are may be disproportionately affected by such PHEICs. This can further compound the intersecting wider inequalities that some sex workers in vulnerable circumstances face, such as homelessness, financial insecurity, domestic violence and social exclusion (Lanau and Matolsci 2024).

Given the significant health inequalities sex workers can face, they should be eligible to receive targeted and tailored health support to reduce health protection risks (Hester 2019; Jeal and Salisbury 2004a). However, they are often not explicitly eligible for targeted and tailored support due to a lack of information on incidence, prevalence of disease, and even more basic data such as reliable estimates of the number of sex workers in the UK. Evidence-based decision making on effective tailored support requires improved understanding of the size and diversity of the sex worker population as well as an understanding of regional variation to guide provision of support services. Improved knowledge would enable health services, vaccination programmes, and other services to better support sex workers and reduce health protection inequalities; both in routine contexts, and during outbreaks or pandemics.

Accordingly, the aim of this paper is to determine a population size estimate, with uncertainty, that is more robust than those currently available. It is acknowledged and recognised, through previous engagement with sex worker communities, that sex workers may have understandable concerns around the motivation of seeking to better quantify the sex worker population. The motivation of this paper is to determine a population size estimate to better inform health decision making to inform improved tailored support and to reduce health inequalities faced by sex workers.

Sex workers are a population who are often underserved by health services, and often missing from existing mainstream health data sets. Experiences of stigma and discrimination, wider legal contexts, and lack of trust due to negative previous experiences can limit disclosure of sex worker status and engagement with surveys (Hester 2019; Lanau and Matolsci 2024). As a result, estimation methods tend to start with the numbers of sex workers engaging with services before attempting to make adjustment for service use (see, for example, Kinnell 1999). These adjustments are highly uncertain, and this uncertainty has not been quantified in previous work. By contrast, the estimate provided by this paper does so.

In this study, we apply Bayesian Evidence Synthesis to bring together historic estimation efforts with recent ONS National Population Estimates and Genito-Urinary Medicine Clinics Attendance Data (GUMCAD) from the UK Health Security Agency (UKHSA). In particular, the parameters that in fact fed into all historic estimates are inherently uncertain. A key feature of our contribution is that it converts these parameters into prior distributions, so that their inherent uncertainty propagates though to our final estimate of the number of sex workers in the UK. This work represents a methodological first step to better understanding the size of the sex worker population. We recognise that it will be critically important to involve sex worker-led organisations and people with lived experience to work towards coproducing community-led methodological approaches in future work.

## 2 Related Work

Since 1990, multiple studies attempt to estimate the number of sex workers in the UK. While resulting in a point estimate, none of these studies provide quantify their uncertainty. In 2019, Hester et al. reviewed these estimates and provided methodological recommendations for future research.

### 2.1 Population Size Estimation for ‘Hidden’ Populations

Population size estimation for groups who are less likely to appear in administrative data sources and surveys has been an active area of methodological development, particularly in the context of HIV prevention and surveillance among key populations.

These populations are often defined as “hidden” populations in the literature. We recognise that the use of terms such as “Hidden” or “Hard-to-reach/engage” in methodological literature on population size estimation is problematic, as it implicitly suggests reluctance from individuals belonging to these groups to come forward. We recognise that, instead, it is systematic and structural barriers to inclusion that mean these groups can be excluded from representation in mainstream data. These barriers include experiences of stigma and discrimination, wider legal contexts and significant challenges in accessing services. Due to the lack of available data, less evidence is available to policy makers to ensure these groups are included in health policies and wider public services such as in the case of sex workers. We choose to use the term ‘Hidden populations’ in this paper solely for consistency with the published literature, and thereby to maximise the chance that this paper benefits the groups in question.

A recent global scoping review by Xu et al (2022) identifies five main estimation approaches, respectively based on independent samples (including capture-recapture and multiplier methods), population counting (including Delphi and mapping methods), official reports (workbook method), social networks (respondent-driven sampling and network scale-up), and data-driven technologies (Bayesian estimation, stochastic simulation, and other advanced statistical approaches).

The capture-recapture method has been widely applied across different contexts, with studies demonstrating its utility in various African settings. In Rwanda, Musengimana et al. (2021) used capture-recapture alongside enumeration and multiplier methods to estimate the size of the female sex worker population. Similarly, research in Zimbabwe employed multiple estimation methods across 20 sites, using respondent-driven sampling surveys combined with service and unique object multipliers, census, and capture-recapture approaches to generate site-level estimates that were then aggregated nationally (Fearon 2021). We recognise that the unfortunate term ‘Capture-recapture’ might feel stigmatising and cause concern. The methodology, and hence its name, originates from the field of ecology and the need to survey wildlife populations to estimate the total population size. We use this term purely for clarity and consistency with existing literature.

The network scale-up method has gained particular attention as a cost-effective approach for hidden population estimation. Studies in Iran carried out by Olfati et al (2023) have demonstrated the application of network scale-up methods with appropriate adjustments for visibility and popularity factors to account for the stigmatised nature of sex work. This method relies on the assumption that the prevalence of behaviour in respondents’ networks can be generalised to the broader population, though it requires careful adjustment for potential biases.

### 2.2 Sex Worker Population Estimation in the UK Context

The most frequently cited UK estimate originates from Hilary Kinnell’s 1999 study for the European Network for HIV/STD Prevention in Prostitution (EUROPAP), which estimated approximately 80,000 sex workers in the UK (Kinnell 1999). However, Kinnell herself acknowledged significant limitations in her methodology, which relied heavily on extrapolation from service provider contacts and included substantial guesswork given the hidden nature of the work.

Building on Kinnell’s foundations, Cusick et al. (2009) conducted a comprehensive analysis titled ‘Wild guesses and conflated meanings? Estimating the size of the sex worker population in Britain.’ This study reported updated counts of sex workers in contact with specialist services across Scotland and England, applying methods and multipliers derived from the Kinnell study to provide updated population estimates. The authors explicitly acknowledged the limits of their estimates and highlighted the ‘theoretical and methodological difficulties’ inherent in estimating hidden population sizes, while arguing that political and media claims about sex work often conflate different phenomena such as trafficking and abuse. This point is also reiterated by Pitcher (2015).

At the ONS, Abramsky and Drew (2014) used a multiplier method to calculate the number of women sex workers in the UK by first calculating an estimate of the women sex workers per capita in London in 2004 and then multiplying this figure by the ONS national population estimate for the same year. The former number was derived from a 2004 study by the charity Eaves, which estimated the number of off-street women sex workers in London, as well as an estimate from the Metropolitan Police of the number of on-street sex workers in London in the same year. Charity or voluntary sector data is consistently lauded by writers as relatively unbiased—service providers have privileged access to a group who tend not to be represented in many public data repositories (Hester 2019). This is often because charities and sex worker-led support organisations have nurtured trusting relationships with sex workers, which is crucial when trying to build better understanding of population size and diversity.

In 2017, ImportIO published an estimate of the total number of sex workers in the UK. This study multiplied the Abramsky and Drew figure by an estimate of the number of men sex workers to yield the total sex worker population in the UK. ImportIO used data from the online platform AdultWork to estimate the man-to-woman sex worker ratio. The study does leverage online platform data, but suffers from several methodological disadvantages.

Brooks-Gordon (2015) produced its estimate of approximately 73,000 UK sex workers by appeal to data from the UKNSWP—the charity now known as the ‘National Ugly Mugs’ (NUMs). The study separates estimates of the total number of sex workers in the UK regions and in London, before summing them. These area-specific estimates were in turn derived by dividing the estimated average number of clients from UKNSWP specialist services in London (or the regions) by the number of specialist services in that area to yield the contacts per service in that area. This figure was then multiplied by the total number of services in that area to yield the estimated total number of sex workers.

The study by Pitcher (2015) attempts to redress limitations with the Kinnell (1999) and Cusick et al (2009) studies by using a mixed-methods approach using two complementary studies conducted between 2009 and 2014. The first is a quantitative survey of service providers aimed to estimate the number of UKNSWP service users by gender, sector, geographical distribution and service needs in England, Scotland and Wales. The second is a qualitative study comprising semi-structured interviews with sex workers aimed at clarifying the nature of sex work experience, working conditions, backgrounds and views on existing policies. The study also involved the use of multipliers for street-based and indoor workers rather than a single multiplier.

The structure of all of these studies is depicted in more depth in the Appendix.

### 2.3 Bayesian Evidence Synthesis Approaches

Bayesian methods are an alternative to standard statistical inference-forms of the frequentist tradition. The Bayesian paradigm can be particularly useful when data is scarce, while simultaneously some pre-existing knowledge (e.g. from literature) exists that can be used to inform the estimates. Such pre-existing knowledge is encoded in the form of ‘prior distributions’ (from the Latin term for ‘previous’) for the parameters in the model, and then updated to ‘posterior distributions’ (from the Latin term for ‘future/following’) leveraging information in the data during the model fitting process.

The application of Bayesian methods to hidden population estimation has emerged as a promising approach for addressing the limitations of single-method studies. Wesson et al. (2018) developed the ‘Anchored Multiplier,’ a novel Bayesian approach that synthesises estimates from multiple multiplier methods while incorporating prior knowledge from literature or stakeholder input. This method addresses the frequently observed problem that multiple estimates using multiplier methods are often discrepant with non-overlapping confidence intervals. Instead, the paper uses a Bayesian framework to recover an estimate from potentially biased data and reduce variability across estimates.

Bayesian evidence synthesis has also been successfully applied to estimate disease prevalence in hard-to-reach populations, as demonstrated in hepatitis C prevalence estimation in New York City, where researchers combined multiple data sources while explicitly accounting for bias and providing estimates with associated uncertainty for high-risk subpopulations including injecting drug users (Tan 2018). The broader epidemiological community has increasingly recognised evidence synthesis methods as essential for epidemic modelling.

### 2.4 Methodological Gaps and Future Directions

Recent UK-focused reviews (Hester 2019; Lanau and Matolsci 2024) have highlighted that no existing single source allows for reliable estimates of the size and characteristics of sex markets. This results in policy being informed by partial pictures. These reviews have specifically recommended local mapping as an underused approach that could inform both policy development and service provision.

Contemporary reviews note that 59% of recent studies employ multiple methods (Xu 2022). This result may reflect a growing recognition that single-method approaches are insufficient for robust population size estimation. Additionally, newer approaches such as successive sampling population size estimation (SS-PSE) using respondent-driven sampling data offer promise for reducing costs and effort while providing estimates that do not rely on separate studies or additional data collection (Johnston 2015; Crawford 2015).

Consultation with sex workers has highlighted understandable concerns from sex workers on disclosure of their sex work status due to a lack of clarity on privacy and confidentiality and how their information will be used. These concerns are also particularly related to the legal framework around sex work in the UK (CPS 2026).

Existing studies tend to exclude migrant sex workers and sex workers belonging to ethnic minorities, those who have dual and plural careers, as well as other ‘hidden’ populations of off-street and online sex workers not attached to services or who do not identify as sex workers. There is no clear best method to estimate the size and diversity of the sex worker population and no single study, including our study, can be used to definitively determine the population size. It is assumed that there is underreporting, that the actual population size is larger, and qualitative information should be used to support any estimates.

Our study contributes to this evolving methodological landscape by applying Bayesian evidence synthesis specifically to the challenge of sex worker population estimation in the UK. In doing so, we address the identified need for approaches that can rigorously combine multiple sources of evidence while appropriately quantifying uncertainty. This work builds particularly on the foundation established by Cusick et al. (2009). It also incorporates advances in Bayesian methodology that have emerged in the subsequent decade and a half.

A recent comprehensive analysis by *Weitzer et al*—currently available only as a pre-print and not yet peer-reviewed—estimated 102,000 sex workers in the UK (range: 75,000-130,000) using a weighted synthesis of four distinct methods: historical averaging, trend extrapolation, sector-based scaling from online platform data, and international comparisons. Our Bayesian evidence synthesis yields a similar central estimate of 84,000 sex workers in the United Kingdom (95% credible interval: 49,000-130,000), but the methodological approaches differ fundamentally. We here say a little about these differences (Weitzer et al. 2025).

Weitzer et al. treat historical studies as black boxes and assign subjective quality weights (5-35%) to combine estimates, whereas our approach explicitly models each study’s data-generating process, treating underlying parameters as uncertain quantities with prior distributions. Their inclusion of a sector-based method that scales Beyond the Gaze online platform data to estimate total market size produces their highest single-method estimate (184,637) but assumes 60-70% of sex workers advertise online—an assumption our model does not make. Both approaches make substantial downward adjustments to the ImportIO (2017) estimate and give considerable weight to practitioner knowledge from organizations like National Ugly Mugs. The convergence of our central estimates (81,000 vs 102,000) despite fundamentally different methodologies provides reassuring triangulation, though the Weitzer et al. findings await formal peer review. However, our Bayesian framework’s key contribution is the formal quantification of uncertainty through posterior distributions with full mathematical specification and convergence diagnostics, rather than deriving confidence ranges from the spread of multiple method results. This methodological rigor makes our approach fully replicable and extensible to incorporate new evidence as it emerges.

## 3 Model structure

### The model employs a three-level hierarchical structure

Level 1 fixes the study-specific estimates. Rather than treating study estimates as black boxes, the model explicitly derives each study’s estimate from its documented parameters (detailed in the Appendix). Each study-specific parameter receives a prior distribution, so that uncertainty in the original methodology propagates through to the derived estimate. The observed study count is then modelled as a noisy log-normal realisation of the model-derived estimate:

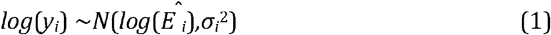

where *y*_*i*_ is the observed count, 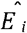 is the mechanistic estimate, and *σ*_*i*_ ^*2*^ = 1/ with *w*_*i*_ as the study quality weight.

Level 2 provides hierarchical pooling via *θ*. Each study’s mechanistic estimate is soft-constrained to a study-specific proportion *θ*_*i*_, scaled by the current UK population:

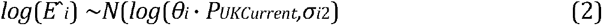

This creates the information pathway from study data to the population-level proportion: study data constrains the mechanistic parameters, the mechanistic estimates constrain *θ*_*i*_, and *θ*_*i*_ constrains *p* through the hierarchical prior.

Level 3 Population-level parameters. Study-specific proportions are drawn from a common beta distribution governed by the overall proportion *p* and between-study heterogeneity *τ*.

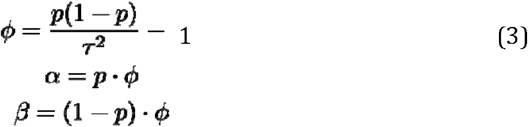

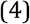

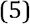

with numerical stability constraints ensuring *α, β* ≥ 0.1. The between-study heterogeneity parameter *τ* captures how much the true underlying proportions vary across different studies, beyond what we would expect from sampling variation alone. The derivation of these equations are set out in more detail in the section 6.1 of the Appendix.

## 4 Evidence to Synthesise

Rather than treating study estimates as black boxes, we explicitly model the data generating process for each study using the documented parameters from the original research papers. We set priors on these parameters and allow uncertainty on them to propagate through to our final estimate. The nature of and relationship between these studies is best understood with reference to the equations and diagram in the appendix.

## 5 Prior Specifications

### 5.0.1 Population proportion

We modelled the prior on the population proportion as follows:

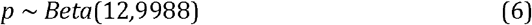

This prior centers the overall proportion at 0.12% with moderate precision, informed by previous research, but allowing the data to influence the inference.

### 5.0.2 Between-study heterogeneity

The parameter *τ* represents between study-heterogeneity. We assign it the following prior:

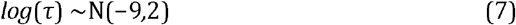

The log-normal parameterisation ensures positive values while allowing reasonable between-study variation. The prior median is exp(-9) ≈ 0.000123.

### 5.1 Quality Weighting System

We used the precedent established by the WHO’s guidelies on estimating the size of populations most at risk to HIV to weight the data sources used as input to our model. The system we built is articulated in more detail in the appendix.

For a given study, 67% of its weighting was assigned on the basis of the data collection method. This was evaluated on the basis of criteria specified in the appendix.

### 5.2 Likelihood Specification

Study estimates are compared to model-derived values using weighted log-normal likelihoods:

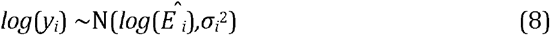

where: 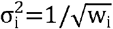
where *y*_*i*_ is the observed study count, 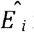 is the model-derived estimate, and 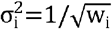. The log-posterior is incremented directly via the normal log-density. The normal density on the log scale ensures symmetry: an estimate *k* times the observed count is penalised identically to one 1/*k* times the observed count. The choice of weighted log-normal likelihoods for comparing study estimates to model-derived values is justified by several considerations, listed in the appendix. The choice of weighted log-normal likelihoods for comparing study estimates to model-derived values is justified by several considerations, listed in the appendix.

### 5.3 Model outputs

The primary output is the total UK sex worker population estimate:

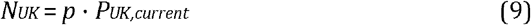

where: *P*_*UK,current*_ = 69,281,437 represents the current UK population.

Secondary outputs include study-specific adjustments, residual analysis, and uncertainty decomposition to assess model fit and identify influential studies.

## 6 Results

### 6.1 Primary Estimates

The Bayesian evidence synthesis model estimated a total of 84,000 sex workers in the United Kingdom (95% credible interval: 49,000-130,000), representing 0.121% of the current UK population. The median estimate was 81,000 indicating a slightly right-skewed posterior distribution. The credible interval spans approximately 89,000 individuals, reflecting a three-fold range from lower to upper bounds.

### 6.2 Parameter estimates and uncertainty

Figure 1 presents the two core model parameters with 95% credible intervals. The dots show the best estimates, while the error bars show the range of plausible values. Longer error bars indicate greater uncertainty. The true proportion estimate (0.122%) carries substantial uncertainty: [0.071%, 0.188%]. The total UK estimate directly scales the proportion to the current population.

**Figure 1.**
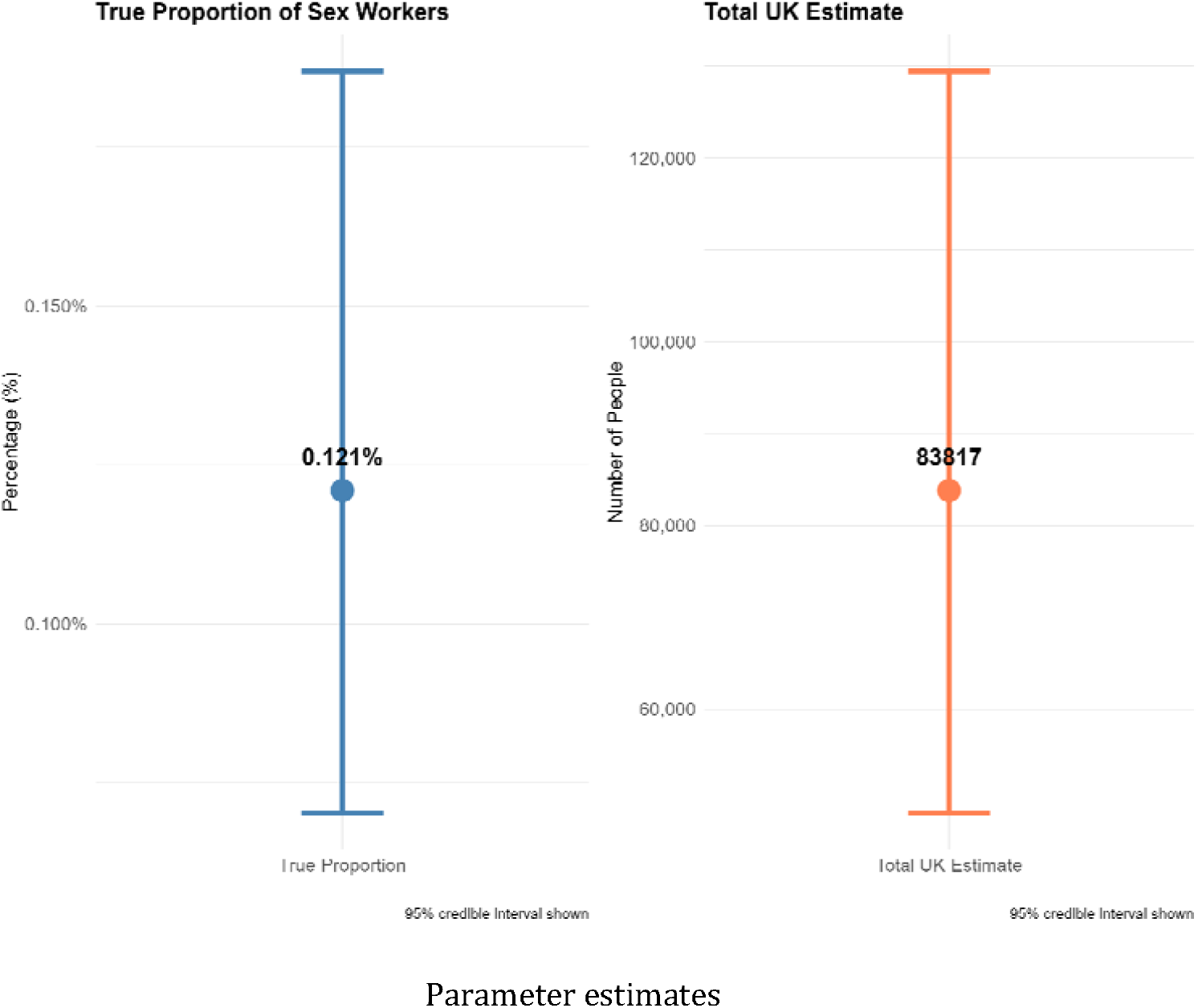
Key Parameter Estimates with 95% Credible Intervals.

**Figure 2.**
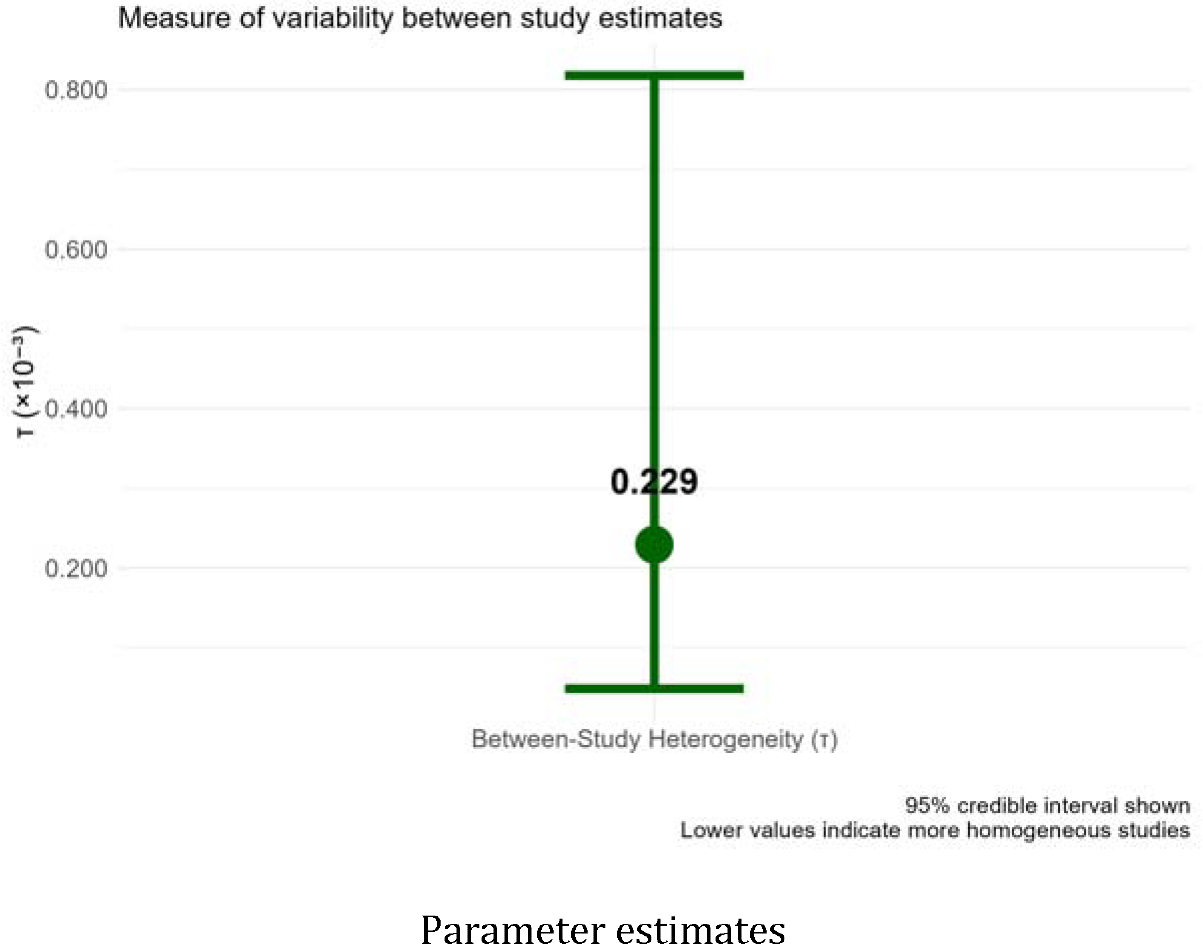
Between-Study Heterogeneity Parameter.

The heterogeneity parameter (tau = 0.000229) indicates moderate between-study variation—studies are neither perfectly consistent nor divergent (Borenstein et al. 2009). This heterogeneity likely reflects genuine methodological differences, temporal changes, and population coverage variations rather than measurement error alone. The relatively modest magnitude suggests the studies are measuring similar underlying phenomena despite some methodological differences. The total UK estimate directly scales the proportion to the current population.

### 6.3 Original vs model-derived estimates

The model made principled adjustments to individual study estimates, motivated by each study’s methodological strengths and limitations. Figure 3 compares original study estimates with model-derived values, with 95% credible intervals.

**Figure 3.**
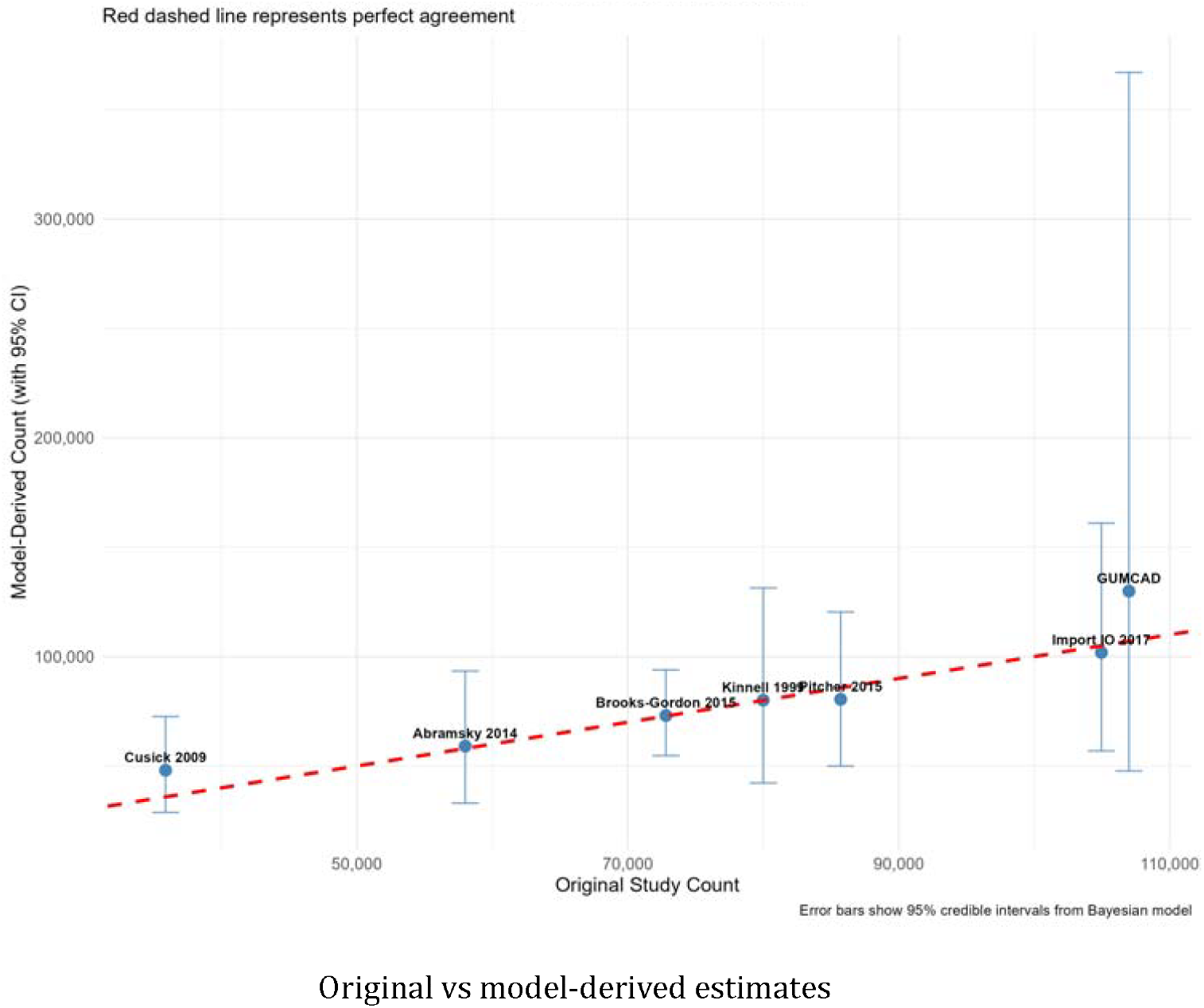
Original Study Estimates vs Model-Derived Estimates.

This scatter plot shows how the model adjusted each study’s original estimate. The red dashed line represents perfect agreement—if all points fell exactly on this line, the model would have made no adjustments. Points above the line indicate the model estimated higher values than the original study; points below indicate downward adjustments. The error bars in Figure 3 represent the uncertainty in each model-derived estimate. Wider error bars indicate greater uncertainty, the result of either lower study quality or inherent methodological limitations.

Key adjustments are shown in Table 2. Three studies—Kinnell 1999, Brooks-Gordon 2015, and Abramsky 2014—sit close to the identity line, indicating that the model’s prior-implied estimates closely reproduce the reported values. Import IO 2017 and Pitcher 2015 are adjusted modestly downward, and Import IO inherits Abramsky’s uncertainty and compounds it with the AdultWork male-to-female ratio. Cusick 2009 is substantially adjusted: its reported count of 35,882 is the lowest in the synthesis, and the model pulls it upward toward the centre, consistent with the prior-implied estimate of approximately 48,000. GUMCAD sits above the identity line. GUMCAD’s credible interval is substantially wider than those of the other studies because its population estimate is derived by dividing the raw count by the product of three rates, each of which carries its own prior uncertainty; small perturbations in any one rate propagate multiplicatively through to the total. Import IO 2017 shows a similarly wide interval for an analogous reason: its estimate compounds Abramsky’s uncertainty with additional uncertainty from the male-to-female ratio.

**Table 1.** Historic Estimates.

| Author | Year | Quantity Estimated | Estimate |
| --- | --- | --- | --- |
| Kinnell | 1999 | Total UK sex workers | 79800 |
| Cusick et al | 2009 | Total UK sex workers | 35882 |
| Abramsky and Drew | 2014 | Women UK sex workers | 60879 |
| Import IO | 2017 | Total UK sex workers | 104964 |
| Pitcher | 2015 | Total UK sex workers | 85714 |
| Brooks-Gordon | 2015 | Total UK sex workers | 72816 |

**Table 2.**
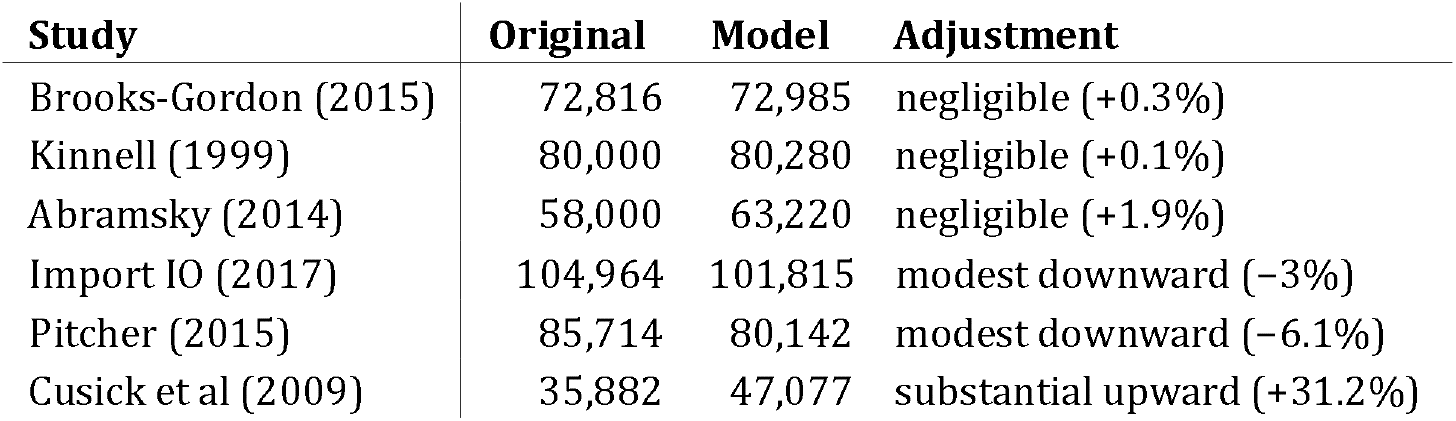
Model adjustments to original study estimates. GUMCAD excluded; see text.

| Study | Original | Model | Adjustment |
| --- | --- | --- | --- |
| Brooks-Gordon (2015) | 72,816 | 72,985 | negligible (+0.3%) |
| Kinnell (1999) | 80,000 | 80,280 | negligible (+0.1%) |
| Abramsky (2014) | 58,000 | 63,220 | negligible (+1.9%) |
| Import IO (2017) | 104,964 | 101,815 | modest downward (−3%) |
| Pitcher (2015) | 85,714 | 80,142 | modest downward (−6.1%) |
| Cusick et al (2009) | 35,882 | 47,077 | substantial upward (+31.2%) |

**Table 3.** Study-specific parameters.

| Variable $w$ | Distribution | First Moment | Second Moment | Reasoning |
| --- | --- | --- | --- | --- |
| $f_{Kinnell}$ | Gamma(11.0,0.0165) | 665.7 | 40364 | Reflects sampling variability |
| $N_{non-resp}$ | Gamma(17.64,8.4) | 2.1 | 0.25 | Relatively low uncertainty |
| $N_{off-street}$ | Gamma(11.0,0.0033) | 3333 | 1010101 | Substantial uncertainty |
| $N_{on-street}$ | Gamma(12.25,0.00175) | 7000 | 4000000 | Moderate uncertainty |
| $S_{regions}$ | Gamma(5.3,0.046) | 115.2 | 2509 | Higher relative uncertainty |
| $a_{Pitcher}$ | Gamma(25,25) | 1.0 | 0.04 | Modest calibration flexibility |
|  | Gamma(30.25,27.5) | 1.1 | 0.04 | Methodological refinement |

Figure 4 displays the percentage residuals; that is, the relative difference between model estimates and original study values. In a sense, the chart shows which studies the model ‘trusted’ most (small bars) versus those it adjusted significantly (large bars). Positive values (blue bars) indicate the model estimated higher than the original study; negative values (coral bars) indicate downward adjustments. GUMCAD is excluded from this plot because the high uncertainty in its adjustment factors produces a residual (31%) that would compress the scale for the remaining studies. Figure 5 explores the relationship between study quality weights and model adjustments. This scatter plot tests whether higher-quality studies (further right on the x-axis) received smaller adjustments from the model (lower on the y-axis). The two studies with the largest adjustments—Cusick 2009 (31%) and GUMCAD (22%)— are adjusted for different reasons: Cusick because its observed count diverges substantially from the prior-implied estimate, and GUMCAD because of compounding uncertainty in the scaling factors used to convert a raw clinical count to a population total. Among the remaining five studies, adjustments are uniformly small (below 7%), regardless of quality weight. The absence of a clear overall trend reflects the fact that adjustments at the study level are primarily driven by the tension between each study’s mechanistic parameter priors and its data likelihood, with the hierarchical constraint through *θ* exerting a secondary influence given the wide uncertainty on individual studies.

**Figure 4.**
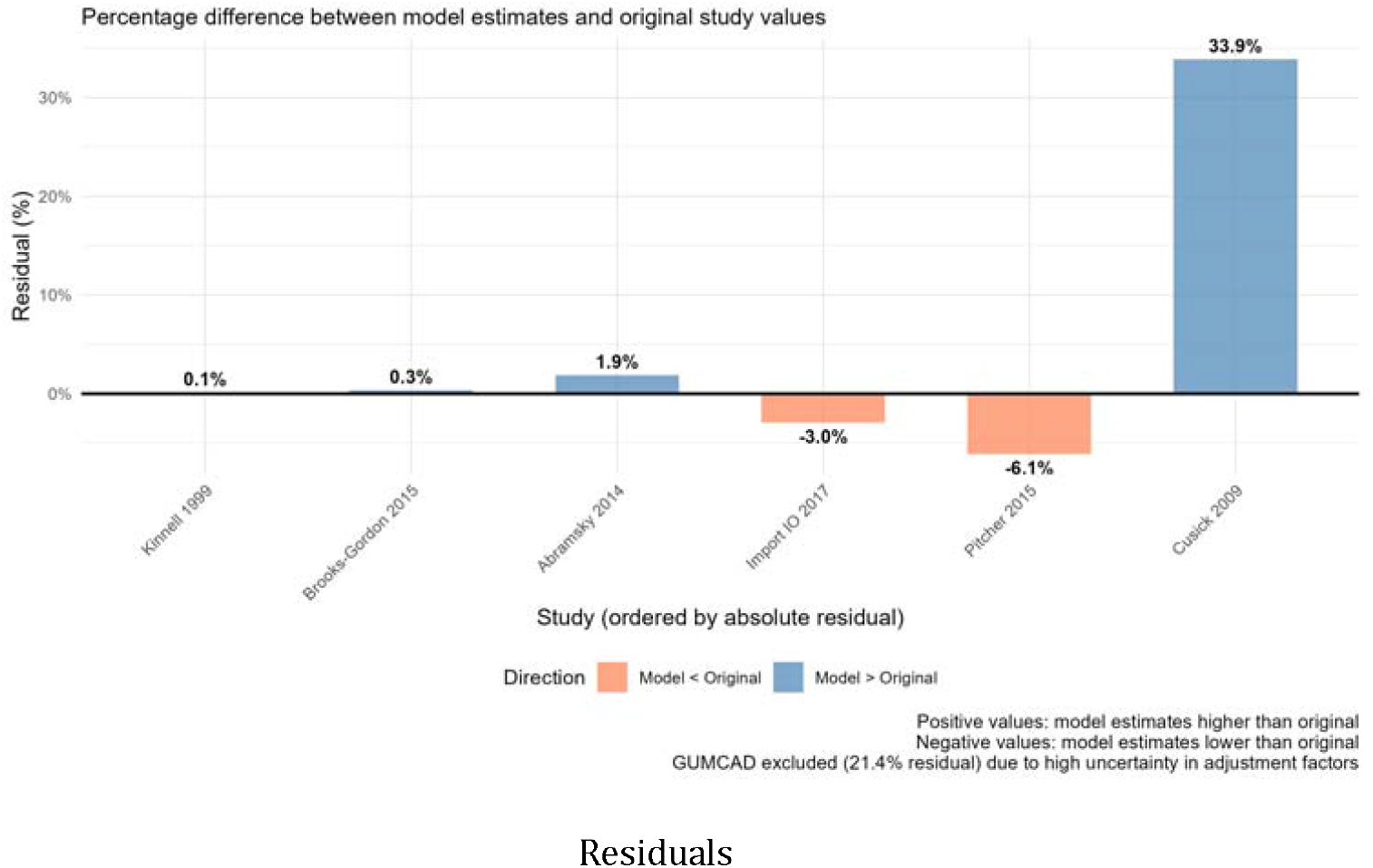
Model Residuals by Study (Excluding GUMCAD)

**Figure 5.**
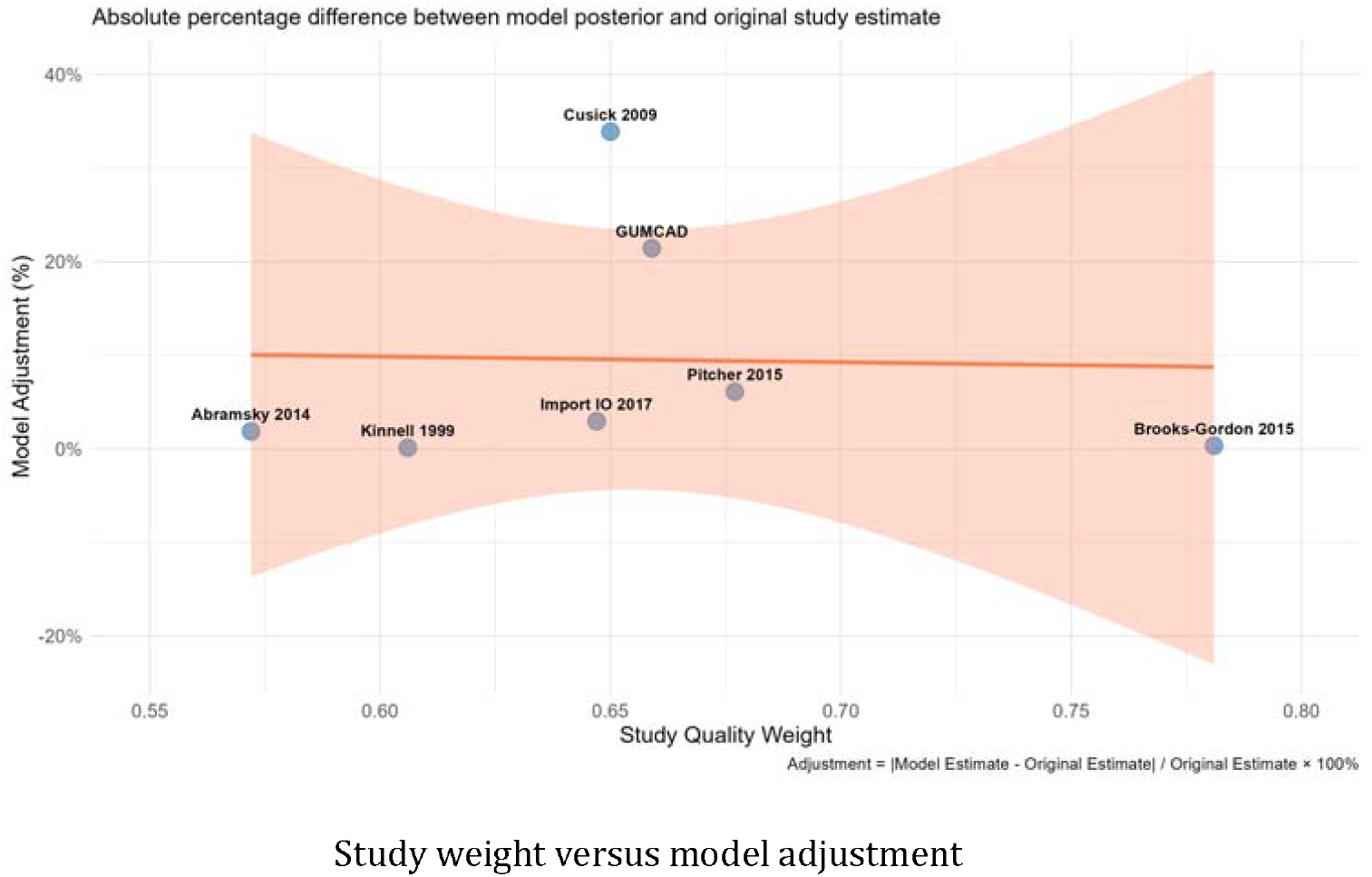
Study Weight vs Model Adjustment.

### 6.4 Sensitivity analyses

We conducted sensitivity analyses to assess model robustness. First, we implemented a leave-one-out analysis that systematically removed each study by setting its quality weight to 0.01 (effectively zero influence) and re-fitting the model. The resulting estimates ranged from 82,633 (GUMCAD removed) to 85,781 (Cusick removed), a span of 3,148 with a coefficient of variation of 1.20%. No single study’s removal shifts the headline estimate by more than 2.3%, confirming that the synthesis is not driven by any individual data source.

Additionally, we compared our quality-weighted approach with uniform weighting (1/N) to assess sensitivity to the weighting scheme. With quality weighting, the central estimate was 83,817; with uniform weighting, 83,957 — a difference of 0.2%. This near-identity confirms that results are robust to the choice of weighting methodology.

The modest sensitivity to both individual studies and weighting specification reflects the structure of the model: the Beta(12, 9988) prior on the population proportion carries considerable prior information, and the seven studies—each with wide individual uncertainty—do not collectively provide sufficient precision to shift the posterior substantially. The robustness demonstrated here is therefore partly a consequence of prior dominance, rather than of strong agreement among the studies.

### 6.5 Posterior predictive performance, and sensitivity to priors

The model successfully reproduced the observed pattern of study estimates, and also accounted for their methodological limitations. All original study estimates fell within the 95% credible intervals of their corresponding model-derived values. This suggests an adequate model fit without over-fitting to any individual study.

The relatively tight agreement between study-specific estimates and the overall synthesis suggests the data dominated the prior specifications. The beta(12, 9988) prior on the overall proportion carried moderate information content but was overwhelmed by the collective evidence from seven studies. Similarly, the gamma priors on study-specific parameters centered the estimates around observed values while allowing data-driven adjustments.

## 7 Discussion

### 7.1 Principal Contributions

This study makes three key contributions to sex worker population estimation in the UK.

First, we provide a revised estimate through comprehensive evidence synthesis. Unlike previous single-method approaches, our Bayesian framework systematically combines seven studies spanning 1999-2023, accounting for methodological differences and study quality. This synthesis approach reduces the influence of any single study’s limitations while leveraging the collective information across all available evidence sources.

Second, we quantify uncertainty in a methodologically principled way. Previous estimates provided point estimates without confidence intervals despite the inherent challenges in estimating hidden populations. Our approach propagates uncertainty through all model parameters—from study-specific multipliers to between-study heterogeneity—yielding credible intervals that reflect our knowledge limitations. The resulting estimate of 84,000 sex workers in the United Kingdom (95% credible interval: 49,000-134,000) provides policymakers with quantified uncertainty essential for robust planning.

Third, we establish an extensible framework for future research. Our hierarchical Bayesian model can readily incorporate new studies, updated population data, or novel estimation approaches without requiring complete re-analysis. As new evidence emerges—whether from surveillance systems, digital platforms, or innovative methodologies—the framework can integrate these sources while maintaining methodological consistency.

### 7.2 Key Limitations

First, the underlying studies lack standard errors, preventing uncertainty-based weighting. We addressed this through systematic quality assessment, but individual study uncertainties remain unquantified. This limitation is a consequence of a broader challenge: most sex work population studies report point estimates without accompanying measures of precision.

Second, our model effectively assumes temporal stability across studies spanning nearly 25 years. The sex work landscape may have evolved significantly due to technological changes (such as online platforms), economic factors, legal developments, or societal attitudes. Without explicit temporal modeling, we cannot determine whether the population is growing, declining, or remaining stable over time.

Third, despite methodological advances, we remain fundamentally constrained by the quality of underlying evidence. Our approach still relies on multiplier methods and extrapolations that inherit the subjective assumptions and coverage limitations of original studies. More rigorous methodologies like capture-recapture or respondent-driven sampling, which rely on individual-level data rather than service-level aggregates, would provide stronger inferential foundations.

Fourth, our approach did not include coproduction with people with lived experience. Future more rigorous methodologies which rely on individual-level data should be co-designed and co-delivered with sex worker led organisations including people with lived experience.

### 7.3 Scale Implications for Public Health Practice

The size of the sex worker population has significant implications for public health intervention design and resource allocation. Having more confidence about the possible scale and range of the estimate allows us to better quantify the health needs of sex workers, identify whether sex workers are missing from services, consider spend per sex worker and help inform more proactive, targeted and tailored approaches to support sex workers and reduce the health inequalities they face. We can also use this estimate to inform public health actions to prevent and respond to outbreaks that may affect sex workers.

The UK Health Security Agency (UKHSA), responsible for providing scientific and operational leadership to protect the public’s health in the UK, published an STI Prioritisation Framework in 2024 that provides an evidence-based approach to local prioritisation for sexual health services which identifies target groups for STI prevention based on health inequalities (GOV.UK 2025). The framework specifically mentions sex workers as one of the inclusion health groups that face greater barriers to accessing sexual health services. Recent research looking at primary and secondary care data (Simms-Williams 2026) found sex workers to be at increased risk of both physical and mental health conditions compared to non sex workers, thus highlighting the urgent need to provide tailored, non-judgmental, and trauma-informed health interventions for sex workers to reduce these inequalities. UKHSA’s operational guidance for STI outbreaks explicitly mentions that voluntary sector organisations can help engage with inclusion health groups including sex workers, recognising the importance of tailored support (GOV.UK 2024).

It is hoped that this estimate will help to inform tailored support and evidence-based decision making to reduce the inequalities experienced by sex workers. For comparison with sizes of other key populations who can face sexual health inequalities, ONS and NATSAL data indicates that men who have sex with men make up around 1% of the UK population. Increasing awareness of HIV testing among those at higher risk which would equate to approximately 670,000 people in a UK population of 67 million (UKHSA 2016). Global estimates suggest people who inject drugs typically represent around 0.2-0.4% of adult populations in developed countries which would suggest there are roughly 134,000-268,000 people who inject drugs in the UK (Degenhardt et al. 2017). Our estimate of approximately 81,000 sex workers (0.121% of population) is notably smaller than those in the population who identify as men who have sex with men (1%) but potentially similar to or larger than the estimated number of people who inject drugs. This suggests sex workers represent a substantial but proportionally smaller group that deserves dedicated public health attention, particularly given UKHSA research suggesting that sex workers can experience higher rates of sexually transmitted infections compared to other populations (UKHSA 2014).

While this analysis provides a crucial national baseline, the nature and extent of sex work varies considerably across geographic areas, sometimes even between neighboring local authorities. Economic factors, transportation networks, regulatory environments, and societal attitudes all influence local sex work markets. A robust modeling framework should ultimately provide geographic breakdowns that can inform local public health planning and resource allocation.

We intend to codevelop our modeling approach to enable subnational estimates that can guide targeted interventions in a way that respects the privacy and safety of local communities. This geographic specificity is particularly important for outbreak response, where understanding local population densities and network structures can inform infection control measures.

Additionally, future work will aim to disaggregate estimates by type of sex work, particularly focusing on forms that present elevated risks for STI transmission and close-contact communicable diseases. Different modes of sex work—street-based, escort services, online platforms, venues-based—involve different risk profiles, service needs, and intervention opportunities. Developing typologically-specific estimates will enable more precise public health planning and more effective harm reduction approaches.

While population size estimation is undertaken here to support health protection and more equitable access to services for sex workers, sex worker-led literature cautions that mapping and enumeration can carry significant risks in criminalised or enforcement-linked contexts when data are used beyond health purposes (NSWP 2015). Wider public health and social research demonstrates that policing and enforcement practices can be associated with harm, and there are risks that data collected for health purposes can be repurposed in ways that increase surveillance and undermine trust (Platt et al. 2018; Grenfell et al. 2023). Future efforts to develop or refine estimates for health purposes should therefore prioritise peer-led, community-governed approaches, with robust safeguards against data repurposing and a clear, demonstrable link to improving sex workers’ health, safety and access to care.

### 7.4 Conclusion

Our analysis provides the most comprehensive and methodologically rigorous estimate of the UK sex worker population available, synthesising nearly two decades of research through sophisticated Bayesian methods. The estimate of approximately 84,000 individuals, with substantial but well-characterised uncertainty, can better inform evidence-based public health planning that embraces rather than ignores epidemiological uncertainty.

Most importantly, these findings underscore the need for continued investment in coproduced approaches to better understand and quantify the size, diversity and health needs of sex worker communities. The population size we identify is substantial enough to warrant dedicated public health attention and resources, while the uncertainty we quantify highlights opportunities for methodological advancement through community engagement and innovative peer-led data collection approaches.

Future progress requires moving toward partnerships methodological approaches with sex worker communities and specialist service providers. Only through such collaboration can we develop both more accurate, inclusive and acceptable population estimates and more effective interventions that genuinely serve the health and wellbeing of these communities and contribute to broader public health objectives.

## Data Availability

All data present in the study comprises of results found in previously published literature, with the exception of GUMCAD data. GUMCAD is a dataset held internally by the UK Health Security Agency (UHKSA). Population figures are from the Office for National Statistics' (ONS) Population Estimates and available on the ONS website.

## Statements and Declarations

We have no relevant financial or non-financial interests to disclose. There are no competing interests to report. All work in the main text is the authors’ own.

## Credit Author Statement

Harry Long: Conceptualisation, Methodology, Formal Analysis, Investigation, Writing - Original Draft, Visualisation;

Leonardo Gada: Methodology, Formal Analysis, Investigation, Writing – Original Draft, Visualisation, Supervision, Project administration;

Lois Murray: Writing - Review and Editing, Conceptualisation;

Timothy Laurence: Supervision, Validation;

Andrew Hayward: Supervision, Validation;

Thomas Finnie: Supervision, Validation.

### 8 Appendix

#### 8.1 Hierarchical model

We here derive equations (3)-(5). Recall that our hierarchical model uses a Beta distribution for the study-specific proportions. However, directly specifying priors on *α* and *β* is problematic for a number of reasons. First, they are not intuitive parameters. Second, they’re constrained (because they must be positive). Third, they don’t directly relate to meaningful quantities like mean and variance. The parameterisation with ⍰ that we elected—a standard “mean-precision” parameterisation—solves these issues. In particular, recall that *p* is just the overall proportion (mean of the Beta distribution), whereas *τ* is the between-study heterogeneity parameter. The key relationships are as follows:

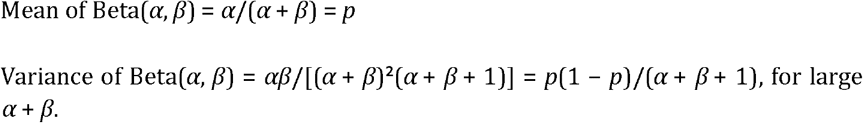

We want to control the variance through *τ*, so we set:

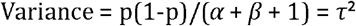

Solving for *(α + β)*: *α + β*

+ 1 = p(1-p)/τ^2^ *α + β* = p(1-p)/*τ*^2^ - 1

This is our ⍰:

⍰ = *α + β* = p(1-p)/*τ*^2^ - 1 [Equation 3]

Then, using the mean constraint, we get:

*α* = p · ⍰ [Equation 4]

*β* = (1-p) · ⍰ [Equation 5]

In sum, the mean-precision parameterization makes the model more interpretable and computationally stable than working directly with *α* and *β*. The proportion p directly represents the overall value we’re estimating. *τ*^2^ effectively controls how much studies vary around *p*. The constraint *α, β* ≥ 0.1 prevents extreme distributions. We use the beta distribution, of course, because it works well with proportion data.

#### 8.2 The structure of the studies

##### 8.2.1 Kinnell 1999

Kinnell’s original estimate is derived in the following simple way:

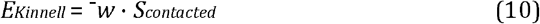

where: *E*_*Kinnell*_ is the Kinnell estimate of the number of UK sex workers, 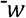 is the average workers per service (665), and *S*_*contacted*_ is the number of sex workers known to be working with support services (120).

##### 8.2.2 Cusick et al 2009

In the Kinnell study, 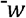 is 2.1x larger than *S*_*contacted*_. Cusick et al incorporate this factor into their estimate:

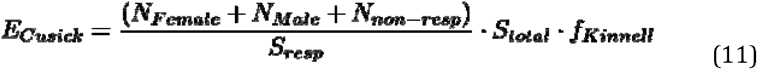

where: *N*_*female*_ is the number of reported female sex workers (11,134), *N*_*male*_ is the number of reported male sex workers (1,493), *N*_*non−resp*_ is the non-responder estimate (parameter 5), *S*_*resp*_ is the number of specialist services that responded (38), *S*_*total*_ is the total number of specialist services (54), and *f*_*Kinnell*_ is the Kinnell adjustment factor (2.1).

##### 8.2.3 Abramsky and Drew 2014

The estimate, produced for the Office of National Statistics, performs a geographic extrapolation from London to the UK:

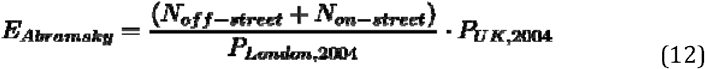

where *N*_*off−street*_ is the number of off-street female workers in London (parameter 6), *N*_*on−street*_ is the number of on-street female workers in London (parameter 7), *P*_*London*, 2004_ is the London population of 2004 (7,389,100), and *P*_*UK, 20*04_ is the UK population of 2004 (59,870,000).

##### 8.2.4 ImportIO Model 2017

This estimate builds on the Abramsky and Drew estimate by using online platform ratios:

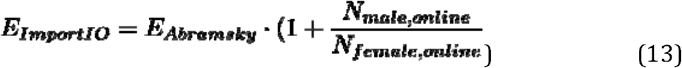

such that: *N*_*male,online*_ is the number of male workers on the online platform AdultWork in London (2,959), and *N*_*female,online*_ is the number of female workers on AdultWork in London (4,086).

##### 8.2.5 GUMCAD

From experts within UKHSA, we received counts of the absolute number of patients using GUMCAD clinics who were also identifying as sex workers for each year between 2011 and 2023 inclusive.

Some deduplication has been applied to these counts. On the one hand, GUMCAD dataset is pseudonymized, and thus names, dates of birth, NHS identification numbers, postcodes etc are not reported. Consequently, we are not able to precisely deduplicate across clinics. On the other hand, patients can be deduplicated if they attend the same clinic multiple times and report different sexual orientation throughout the year. Thus, our data excludes duplicate patients from the same clinics, presenting them just once per year, based on their sexual orientation with the highest risk.

The average of the absolute number of sex worker patients seen per clinic over 12 years between 2011 and 2023 is approximately 3,700. Since these counts were solely for England, we multiplied this figure by 1.19 to yield 4,414. Thus, 4,414 is our absolute count that we input into our model. Of course, we need to adjust for ascertainment bias here. We model this bias with two factors:

1. Rate seeking healthcare: the proportion of sex workers who seek healthcare;
2. Identity disclosure rate: the proportion of sex workers who disclose theiridentity as sex workers.

A recent study suggests a 38% identity disclosure rate (Jeal and Salisbury 2004b). We adjust this to 25%, on the basis that Bristol is a much more accepting city of sex work than the UK average (thus leading to higher disclosure rates than would be representative of the UK average).

For the rate seeking healthcare factor, we elect 33% (Jeal and Salisbury 2004b). Using these factors to calculate the total number of sex workers, from the GUMCAD count of 4414, would run as follows. Let x stand for the total number of sex workers in the UK. Then:

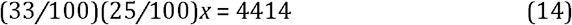

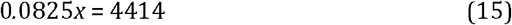

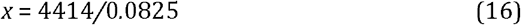

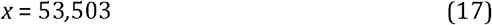

We set priors on these factors. The healthcare seeking rate was centred at 33% with tight uncertainty (although this could reasonably be expanded in accordance with further expert testimony):

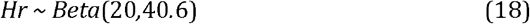

To center it at 25% with moderate uncertainty, we gave the disclosure rate the following prior:

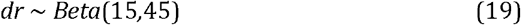

##### 8.2.6 Brooks-Gordon 2015

The Brooks-Gordon estimate combines regional and London estimates:

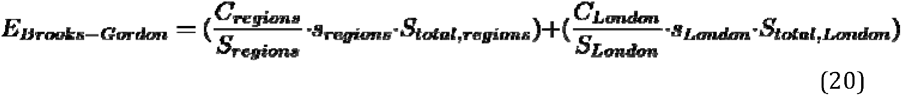

where:

*C*_*regions*_,*C*_*London*_ are the number of clients in regions (5,249) and London (3,199) respectively;

*S*_*regions*_,*S*_*London*_ are the numbers of services providing data in the regions (18) and London (4) respectively;

*S*_*total,regions*_, *S*_*total,London*_ are the total numbers of services in the regions (140) and London (40) respectively; *s*_*regions*_, *s*_*London*_ are scaling parameters for methodological adjustment.

##### 8.2.7 Pitcher 2015

Pitcher uses a stratified multiplier approach where different sectors receive different scaling factors based on the observed relationship between service users and estimated populations in each sector. Due to its similarity to the Brooks-Gordon method, we model the Pitcher estimate as an adjustment of Brooks-Gordon:

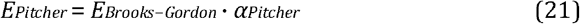

#### 8.3 Likelihood Specification

Study estimates are compared to model-derived values using weighted log-normal likelihoods. We here justify such likelihoods for such a purpose.

First, log-normal distributions have support on (0, inf), naturally respecting the constraint that population size estimates must be positive. Taking the log transforms the problem to the real line where normal distributions are appropriate. Second, estimation errors are likely multiplicative rather than additive. For instance, a study estimating 50,000 might be off by a factor of 1.5-2x; whereas a study estimating 500,000 might be off by the same factor, not the same absolute amount. Log-normal captures this: log(observed) = log(true) + error, where error is normally distributed.

Third, population size estimates tend to have right-skewed uncertainty, in that they are more likely to underestimate than overestimate (due to hidden populations). Confidence intervals are often <u>asy</u>mmetric (e.g., 50,000-150,000, not 75,000 ± 25,000). Log-normal naturally produces this desired asymmetry. Fourth, our weighting scheme . Consequently, higher quality studies (larger *w*_*i*_) will therefore possess smaller variance (smaller *σ*_*i*_^2^). Lower quality studies will possess larger variance. In accordance with intuition, this down weights unreliable estimates in the likelihood.

Finally, log-normal likelihoods are well-behaved in MCMC sampling. With truncated normal distributions, by contrast, common implementations can run up against boundary issues. Standard optimization routines handle log-normal efficiently.

#### 8.4 Study-specific parameters

All positive study-specific parameters use gamma priors (to avoid boundary issues):

Alternatives to gamma distributions suffered from shortcomings. For example, if we had used normal priors, there would be non-zero probability mass on negative values. A truncated normal would be more promising, but creates computational complications at the boundaries. A uniform distribution would not incorporate our prior knowledge about reasonable parameter ranges.

#### 8.5 Detailed account of the Evidence Quality Weighting System

The criteria used to evaluate the study weighting system run as follows:

##### 1 Population Coverage (67%)

- Are all sectors of sex work represented in the sample?
  – Bar-based or hostess bars
  – BDSM, kink and fetish
  – Brothels, massage parlours and saunas operating as brothels
  – Erotic massage
  – Erotic and exotic dance
  – Escorting
  – Pornography, glamour and erotica
  – Sex parties
  – Street/outdoor
  – Sugar arrangements
  – Telephone, text-based, TV-based, live voyeurism
  – Therapeutic services
  – Webcamming

- Are all genders represented in the sample?
  – Cis Men
  – Trans Men
  – Cis Women
  – Trans Women
  – Other Genders

- Are all age groups represented in the sample?
  – 16-24
  – 25-34
  – 35-54
  – 55+

– Are all geographic localities represented in the sample?
  – North East
  – North West
  – Yorkshire and Humber
  – East Midlands
  – West Midlands
  – East of England
  – London
  – South East
  – South West

– How different is the target population at the time of the study from the target population in the present day (2026)?
– Is there evidence of a more general mechanism that prevents the sample in the data source from being the target population?

##### 2 Method-specific criteria (33%)

- Multiplier Method
  – Are the two data sources (survey and multiplier) independent?
  – Is the survey random, and does it include the group described in the multiplier?
  – Are the population in the survey and multiplier defined in the same way?
  – If multiple sources are used, are time periods, age ranges and geographic area aligned?
- Population Enumeration/Census
  – What is the extent of the community guides’ outreach? Did they cover small areas? data collected from different regions and establishments vary in important ways?
  – Did the method capture individuals beyond the strictly defined population?

To foreshadow what’s to come, all weighting schemes contain ineliminable subjective elements and require intuitive judgements at some stage in the justificatory chain. Nonetheless, the judgements we have made are:

1. supported by principles of sampling theory, hidden population methodology,and evidence synthesis
2. empirically calibrated against known cases (Johnston 2015)
3. mathematically coherent with sensible consequences for edge cases
4. precedent-based, drawing on GRADE, Cochrane, WHO guidelines

Where reasonable people may disagree (*e.g*., 85% rather than 80% for PC weight) the system’s outputs change minimally; relative study rankings would likely be identical, for instance, with such changes to the *PC* parameter.

##### 8.5.1 The Overall formula

Our overall weighting formula is as follows:

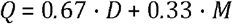

where *Q* = the study’s overall quality weight, *D* = the data collection method, *M* = the method-specific criteria.

Data collection method *D* receives the dominant weight (67%) because it determines who can possibly be included in a study. Even perfect execution of a flawed methodology cannot overcome data quality issues and bias, such as systematic coverage gaps.

There is good empirical support for this choice. The Johnston et al. (2015) formative research paper demonstrates that inappropriate sampling methods lead to complete survey failure (Podgorica FSWs: 1 participant; St Vincent MSM: no recruitment chains). By contrast, method-specific execution issues (like suboptimal multipliers) can be corrected or accounted for analytically.

There is also considerable precedent for assigning *D* the dominant weight in the formula for *Q*. The GRADE system for evidence quality similarly prioritises study design over other quality dimensions, and assigns randomised trials initially high quality regardless of execution details, which are then adjusted downward (Guyatt 2008).

The choice to assign *D* the greatest weight was compared to alternative choices. For instance, equal weighting (50% each) was rejected, because it treats fundamental design flaws equivalently to correctable execution issues. The 2:1 ratio (67%:33%) appropriately reflects that coverage and representativeness are roughly twice as important as technical execution quality.

Method-specific criteria assess whether a chosen method is properly executed, which matters substantially, but is secondary to whether the right method was chosen; it is thus accorded 33% weight. A perfectly executed multiplier method with non-independent sources, for instance, is less problematic—*ceteris paribus*—than a method that excludes 70% of the target population.

The balance gained is intuitively appropriate. For instance, a study with good population coverage (high *D*) but poor technical execution (low *M*) receives 67% weight (0.67 · 100 + 0.33 · 50), while a study with poor coverage (low *D*) but perfect execution (high *M*) receives 50% weight (0.67 · 50 + 0.33 · 100).

There is also considerable precedent for this choice. Meta-analysis guidelines (Cochrane Handbook) similarly treat within-method execution as important, but secondary, to fundamental study validity.

We have chosen to omit uncertainty quantification from the formula for Q for several reasons. First, none of the historical UK studies incorporated uncertainty into their methodologies or reported confidence intervals. For this reason, including a naïve measure of uncertainty as a criterion would penalize all existing studies equally, and thus provide no discrimination between them. It would therefore be rationally eliminable. Additionally, expert stakeholder consultation indicated that uncertainty quantification, while desirable for future research, should not be retroactively applied as a quality criterion to historical estimates that were conducted under different methodological norms.

##### 8.5.2 Formula for D

The formula for data collection method D is:

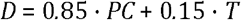

We assign population coverage *PC* 85% weight because population coverage is the dominant component of data collection quality in the context of population size estimation. It determines representativeness. Intuitively, a study from 2010 that captured all sex worker demographics (*PC*=100) is more valuable than a 2024 study of only online female sex workers (*PC*=40), even though the latter is more recent.

There is also considerable empirical support for assigning such high weight to whether a given estimate has representative population coverage:

1. The Weitzer et al (2025) pre-print’s sector-based method (using ‘Beyondthe Gaze’ online data, scaled up) received substantial downward adjustment precisely because of coverage limitations, despite being recent;
2. Cusick et al (2009) stress that the core problem in sex worker estimationis who is being counted.

Assigning *PC* such a high weight can be given plausible mathematical justification. In particular, with 85% weight, a study with perfect coverage (*PC*=100) that is nonetheless 15 years old (*T*=86) receives *D*=99.9. A study with poor coverage (*PC*=40) but current (*T*=100) receives *D*=49. Intuitively, this correctly prioritises coverage while still valuing recency. Moreover, the weight we assign to temporal relevance warns against blindly quoting or re-using estimates that are decades old, for such usage implictly assumes that the UK’s population and nature are static.

We assign temporal relevance *T* the relatively modest weight of 15% because while sex work markets do change over time (particularly with technology shifts toward online platforms) core population dynamics change more slowly than, say, drug markets or disease prevalence. This choice has been empirically calibrated:

1. 15% weight means a 10-year-old study loses 6.3 points (15% *× 42 point lo*ss from T decay)
2. A 20-year-old study loses 11.4 points
3. Intuitively these consequences are approximately right: Kinnell 1999 shouldn’tbe weighted equally to Brooks-Gordon 2015, but the age difference alone shouldn’t dominate if Kinnell had better coverage.

We did consider alternatives to this weighting of temporal relevance. 25% temporal weight was rejected because it would make a comprehensive 2000 study (*PC*=100, T=60) score worse (*D* = 0.85 · 100 + 0.25 · 60 = 100) than a limited 2024 study (*PC*=60, T=100) score (*D* = 0.85 · 60 + 0.25 · 100 = 76). This seems wrong.

Expert judgement in the literature supports our assignment to temporal relevance. For example, *Sanders et al* (Sanders et al. 2017) Beyond the Gaze found online sex work became ‘dominant’ by 2015-2017, but that the transition to this state of the market was relatively slow.

##### 8.5.3 Formula for PC

Consider now our formula for the Population coverage (*PC*) parameter:

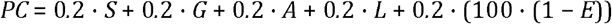

where *S* = sector coverage, *G* = gender coverage, *A* = age coverage, *L* = geographic coverage, and *E* = exclusion mechanisms.

By assigning the five parameters that figure in the formula for *PC* equal weights, we have opted for a uniform distribution.

There are several considerations in favour of a uniform distribution here. Equal weighting treats each dimension as equally important for achieving representative coverage.

This is defensible because equal weighting is a natural consequence of two facts. First, that each dimension represents an independent and equally important axis of potential bias in the estimate. Second, that missing coverage on any single dimension creates selection bias that cannot be compensated for (or even mitigated) by complete coverage on other dimensions. To assign non-uniform weight would be to make an implicit judgement that, say, age mattered more than gender in our estimate. But any such judgement seems quite odd.

It is important to express why certain alternatives were rejected. First, consider alternative, which we have not pursued, of weighting by importance or ‘heterogeneity’. The proposal here is to weight dimensions by how much variation they contain in the population. But this strategy confuses prevalence with representativeness.

Consider, for instance,

1. Cis women are ≈ 73.5% of UK sex workers (Beyond the Gaze)
2. Should we weight gender coverage at 15% because ‘most are women anyway’?
3. No: excluding 26.5% of the population (all men, trans people, non-binarypeople) creates systematic bias regardless of their relative size.

Consider now the alternative (which we have not pursued) of weighting dimensions by their correlation with key outcomes (health risks, service access, etc). The worry with this proposal is that we don’t reliably know which dimensions correlate most strongly with outcomes—in fact, that’s research we’re trying to enable with these estimates! Also, this strategy creates a problematic circularity: using assumed outcome patterns to weight studies meant to reveal outcome patterns. It is also true that different stakeholders care disproportionately about different outcomes (HIV risk vs. violence vs. economic security); and so it would be difficult to produce a single weighting scheme serving all uses and about which we would have consensus.

A uniform distribution can also be justified mathematically. Since our modelling approach is Bayesian, here equal weights represent the maximum entropy (minimum assumption) prior when we lack strong evidence for differential weighting. This choice therefore is the mathematical encoding of our present ignorance. Given five independent coverage dimensions, with no principled basis for ranking their importance, equal weighting minimises the information we’re imposing on the system. Any other weighting scheme requires justifying why dimension *X* matters more than dimension *Y* . We may also appeal to a burden of proof principle that applies asymmetrically to non-uniform weights: unequal weights require positive justification, but equal weights are the null hypothesis.

Why (1−*E*) formulation for the exclusion mechanism? The exclusion mechanism uses (1−*E*) so that presence of exclusion reduces the score. The mathematical form ensures:

1. No exclusion (*E* = 0) contributes 100 points
2. Partial exclusion (*E* = 0.5) contributes 50 points
3. Strong exclusion (*E* = 1) contributes 0 points

This is multiplicative with the other coverage components (since we multiply by 1 − *E*), which is intuitively correct: if a study systematically excludes 50% of the population, it doesn’t matter if it covers all sectors/genders/ages among the accessible 50%—coverage is still only half.

Take for example a study covering all demographics (*S* = *G* = *A* = *L* = 100) but with strong evidence of systematic exclusion. (*E* = 1) would receive *PC* = 80 (not 100); making it correctly penalised.

Regarding the sector coverage parameter *S*, we have 13 Sectors because we have followed the established Beyond the Gaze typology (Sanders et al. 2017)—and UK-specific literature (Hester 2019), (Pitcher 2015). The next stage of our research seeks to define the most appropriate categorisation for our public health purposes, so we will revisit this issue.

It may not initially be clear why we have 13 sectors, rather than fewer or more. But consider that:

1. Using fewer categories (e.g., just ‘online’ vs ‘offline’) masks importantheterogeneity. Online escorting □= webcamming in terms of STI risks, working patterns, service access.
2. Using more categories (e.g., subdividing escorts by price point) createsmeasurement issues since studies rarely provide that granularity.

Why do we assign equal weights (a uniform distribution) across all sectors?

We don’t know the relative size of each sector in the UK, and weighting by assumed size would introduce arbitrary assumptions. Equal weighting is the maximum entropy (least assumption-laden) approach.

We did consider weighting by estimated prevalence (e.g., if online escorting is 30% of market, weight it 30%). Rejected because:

1. We don’t reliably know relative sector sizes (that’s partly what we’re tryingto estimate!)
2. This would create circularity: using prevalence assumptions to weightstudies that estimate prevalence.

Relatedly, Beyond the Gaze found online work is ‘dominant’ but represents multiple sectors (escorting, webcamming, etc.).

We opt for 5 categories for gender coverage, because we follow Beyond the Gaze’s finding of:

1. 73.5% cis women
2. 19.4% cis men
3. 3% trans women
4. 3% trans men
5. 2.9% non-binary/intersex

Which suggests that all five categories have measurable presence in UK sex work.

We assign equal weight across genders for similar reasons to those for which we assigned equal weight across sectors. We don’t want to weight by prevalence because:

1. We’re trying to estimate total population including all genders
2. Weighting by prevalence would systematically undervalue studies thatcapture smaller groups
3. Equal weighting ensures studies can’t ignore 19.4% of the population (cismen) with minimal penalty

Regarding age coverage, we used four bands with roughly the following rationale:

1. 16-24: Entry age range, often distinct working patterns and risks
2. 25-34: Peak working years for many
3. Experienced workers, different health/economic profiles
4. Older workers, often overlooked in research

As with other parameters, we assign a uniform distribution to avoid arbitrary assumptions about the relative importance of the categories.

Regarding geographic coverage, we used the standard nine regions of England recognised by the Office of National Statistics. These are:

1. Administratively meaningful for public health planning
2. Capture urban/rural variation
3. Reflect different legal frameworks (Northern Ireland has different sex worklaws)
4. Standard for UK health data reporting

Our equal weighting of regions entails that a London-only study receives L=11% (1/9 regions). This appropriately penalises the study despite London’s disproportionate share of sex workers (likely 25-30% of the UK total) because the goal is not proportional population capture, but rather representativeness across diverse regional contexts. Sex work in London is likely to differ from sex work in Newcastle, Cardiff, or rural Scotland in terms of (but not only of): legal enforcement patterns, cost of living and pricing structures, client demographics, availability of support services, and cultural attitudes toward sex work. A London-only study cannot speak to these regional variations, and therefore cannot support national-level inference regardless of how many workers it captures. Equal regional weighting reflects our priority of geographic comprehensiveness over population-proportional coverage.

Our temporal relevance parameter: *T* = 100 · *exp*(−0.01 · *years*_s_*ince*_s_*tudy*) We have chosen exponential decay here. We say a little about why.

Exponential decay reflects compound change—each year’s changes build on previous years. A 10-year-old study isn’t just 10% worse than a 5-year-old study; it’s missing 5 additional years of compounded change.

Why did we select a −0.01 decay rate? Because such a rate yields intuitively appropriate penalisation of older studies. Witness:

1. 1 year old:*T* = 100 ⍰ *exp*(−0.01 ⍰ 1) = 99.0 (1% penalty)
2. 5 years old:*T* = 100 ⍰ *exp*(−0.01 ⍰ 5) = 95.1 (5% penalty)
3. 10 years old:*T* = 100 ⍰ *exp*(−0.01 ⍰ 10) = 90.5 (10% penalty)
4. 15 years old:*T* = 100 ⍰ *exp*(−0.01 ⍰ 15) = 86.1 (14% penalty)
5. 20 years old:*T* = 100 ⍰ *exp*(−0.01 ⍰ 20) = 81.9 (18% penalty)
6. 25 years old:*T* = 100 ⍰ *exp*(−0.01 ⍰ 25) = 77.9 (22% penalty)

There are also empirical anchors to this choice. A major structural shift to online platforms occurred ⍰ 2010-2015 (Sanders et al. 2017). A study from 2010 (15 years old) should be penalised ⍰ 14%, which this formula delivers.

Consider an alternative we rejected *T* = 100−(*yearssincestudy*⍰2) would give % penalty per year. This strategy was rejected because, inter alia, linear decay implies constant rate of change, but technological or social changes compound. The shift from 2000 to 2010 (pre-smartphone, limited online platforms) was likely slower than 2010 to 2020 (ubiquitous smartphones, multiple platforms, COVID impacts).

Exponential decay captures the fact that early years lose value slowly (1 year = 1% penalty) but old studies degrade faster (year 20 − − > 21 loses more than year 1 − − > 2).

##### 8.5.4 Method-specific criteria First, consider multiplier methods

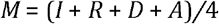

Each criterion represents a distinct violation of multiplier method assumptions: independence (I), randomness (R), definition alignment (D), and parameter alignment (A). Why did we assign equal weight (a uniform distribution) for the four criteria?

Start with independence: if sources aren’t independent, one is double-counting, and the results are to that extent jeopardised. Regarding randomnness, if the survey isn’t random or inclusive, the multiplier doesn’t apply to whole population of interest. On definition alignment, if populations are defined differently, the numbers multiplied together do not represent the populations of interest. Regarding parameter alignment, if time/age/geography don’t align, populations don’t overlap these are logically distinct failure modes with no theoretical reason to weight one more heavily.

These are logically distinct failure modes with no theoretical reason to weight one more heavily. A multiplier study failing completely on one dimension (*I* = 0, others=100) receives *M* = 75, appropriately high but still penalised. Failing on two dimensions (*I* = *R* = 0,*D* = *A* = 100) receives *M* = 50, appropriately middle-range.

Why these specific scoring scales (0/25/50/75/100)? Five-point scales (0/25/50/75/100) provide meaningful gradation, without false precision.

For independence (I):

- 100 = fully independent: Survey and multiplier from completely separate sources (e.g., population survey + NGO service data)
- 75 = largely independent: mostly separate, but minor overlap (e.g., survey includes some NGO clients but randomly selected).
- 50 = partially independent/unclear; paper doesn’t clearly document independence.
- 25 = questionable independence: substantial overlap suspected. 0 = not independent: the same population sampled twice. This maps to Cohen’s (Cohen 1960)—”small/medium/large effect” heuristics adapted for quality assessment.

For randomness (R):

- 100 = random, clearly inclusive: probability sampling that includes the multiplier group.
- 75 = mostly random/inclusive: Minor deviations but fundamentally sound
- 50 = partially random/unclear: Convenience sampling among relevant population
- 25 = minimal randomness: Snowball or venue-based without random recruitment
- 0 = not random/not inclusive: Purposive sampling excluding multiplier group

A justification for this is that random sampling isn’t binary—respondent-driven sampling (RDS) is “semi-random” (Johnston 2010), deserving ⍰ 75. Convenience sampling that’s systematic deserves ⍰ 50.

Consider now enumeration methods:

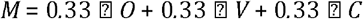

We opt for the uniform distribution because, again, each criterion represents distinct dimensions of enumeration quality:

1. Outreach extent O: was everyone found?
2. Variation handling V: was difference across areas accounted for?
3. Capture precision C: Was only the target population counted?

These are independent failure modes; a study can have comprehensive outreach (*O* = 100) but poor capture precision (*C* = 50) if overcounting, or vice versa.

One alternative we considered (but ultimately rejected) ran as follows: weight outreach extent more heavily (e.g., 50%) since it’s most fundamental. This was rejected because overcapture (*C* = 0) or ignoring regional variation (*V* = 0) equally invalidate estimates.

We also wanted to avoid double-counting: if a study misses street-based sex workers (affects *S*) who are predominantly cis women in certain age groups (affects *G* and *A*), it may appear that we are penalising three times for one problem. This appearance, though, is mistaken, because the dimensions measure different types of exclusion:

- Sector (*S*): Organisational/working pattern exclusion
- Gender (*G*): identity based exclusion
- Age (*A*): age-based exclusion
- George (*L*): place-based exclusion
- Exclusion mechanism (*E*): systematic barrier exclusion

A study could cover all sectors but miss trans people (*S* = 100, *G* = 60) or cover all genders but only urban areas (*G* = 100,*L* = 56); or cover everything but have systematic exclusion of migrants (*S* = *G* = *A* = *L* = 100; *E* = 0.5 ⍰ *PC* = 80.

These are all logically independent axes of coverage.

Take three example hypothetical studies to see the weighting system in action. Let Study A contain perfect coverage, perfect methods, but be relatively old (*PC* = 100,*T* = 81.9,*M* = 100). Then:

- *D* = 0.85(100) + 0.15(81.9) = 97.3
- *Q* = 0.67(97.3) + 0.33(100) = 98.2

Let study B contain limited coverage, good methods, recent (*PC* = 50,*T* = 100,*M* = 100). Then:

- *D* = 0.85(50) + 0.15(100) = 57.5
- *Q* = 0.67(57.5) + 0.33(100) = 71.5

Let study C contain perfect coverage, poor methods, recent (*PC* = 100,*T* = 100,*M* = 25). Then:

- *D* = 0.85(100) + 0.15(100) = 100
- *Q* = 0.67(100) + 0.33(25) = 75.3

This ordering (*A* > *C* > *B*) is defensible. A comprehensive old study with perfect methods (A) beats a recent study with perfect coverage but poor methods (C), which beats a recent study with good methods but limited coverage (B). This appropriately prioritises coverage over recency, and values methodological soundness substantially.

Certain principles pervade the methodology throughout.

Equal weighting as default: Unless there’s strong reason to weight dimensions differently, equal weights minimize arbitrary assumptions

Coverage dominates execution: Who you count matters more than how precisely you count them

Five-point scales (0/25/50/75/100): Provide meaningful gradation without false precision

Exponential over linear decay: Reflects compound change over time

Multiplicative for exclusion: Systematic exclusion invalidates coverage across other dimensions

Independence of dimensions: Each penalty reflects distinct methodological issue, not double-counting

#### 8.6 Robustness

##### 8.6.1 Computational Implementation

The model was implemented in Stan 2.34 using No-U-Turn Sampling (NUTS) with: 4 chains of 20,000 iterations each; 10,000 warmup iterations per chain; adapt delta = 0.999 for numerical stability max treedepth = 15 for complex posterior geometry.

Convergence was assessed using 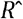 statistics and effective sample sizes, with 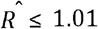, and *n*_*eff*_ ≥ 400 required for all parameters.

##### 8.6.2 Trace Plots

Figure 6 displays trace plots for three key parameters across all four chains. Think of these plots as showing the “path” each chain took while exploring possible parameter values. Good convergence appears in four properties. First, a ‘fuzzy caterpillar’ appearance; the traces look like overlapping, hairy caterpillars with no obvious trends or patterns. Second, there is chain overlap: all four coloured lines explore the same range of values, and intermingle freely. Third, there is no systematic drift, in that the lines don’t consistently increase or decrease over time. Finally, there is stable mixing: the chains move freely around the parameter space without getting ‘stuck’.

Our trace plots demonstrate good behavior. All chains overlap extensively, show no systematic trends, and exhibit the characteristic “white noise” appearance that indicates thorough exploration of the posterior distribution. The true proportion parameter shows particularly clean mixing, while tau (heterogeneity) displays slightly more autocorrelation (but still acceptable behavior).

##### 8.6.3 Model Convergence and Computational Performance

All model parameters demonstrated excellent convergence across the four independent Markov chains. The potential scale reduction factor 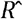 ranged from 0.999 to 1.002 for all parameters, well below the conventional threshold of 1.01 that indicates convergence concerns. The effective sample sizes ranged from 2,731 to 44,198 samples. This substantially exceeds the minimum recommended threshold of 400 effective samples per parameter.

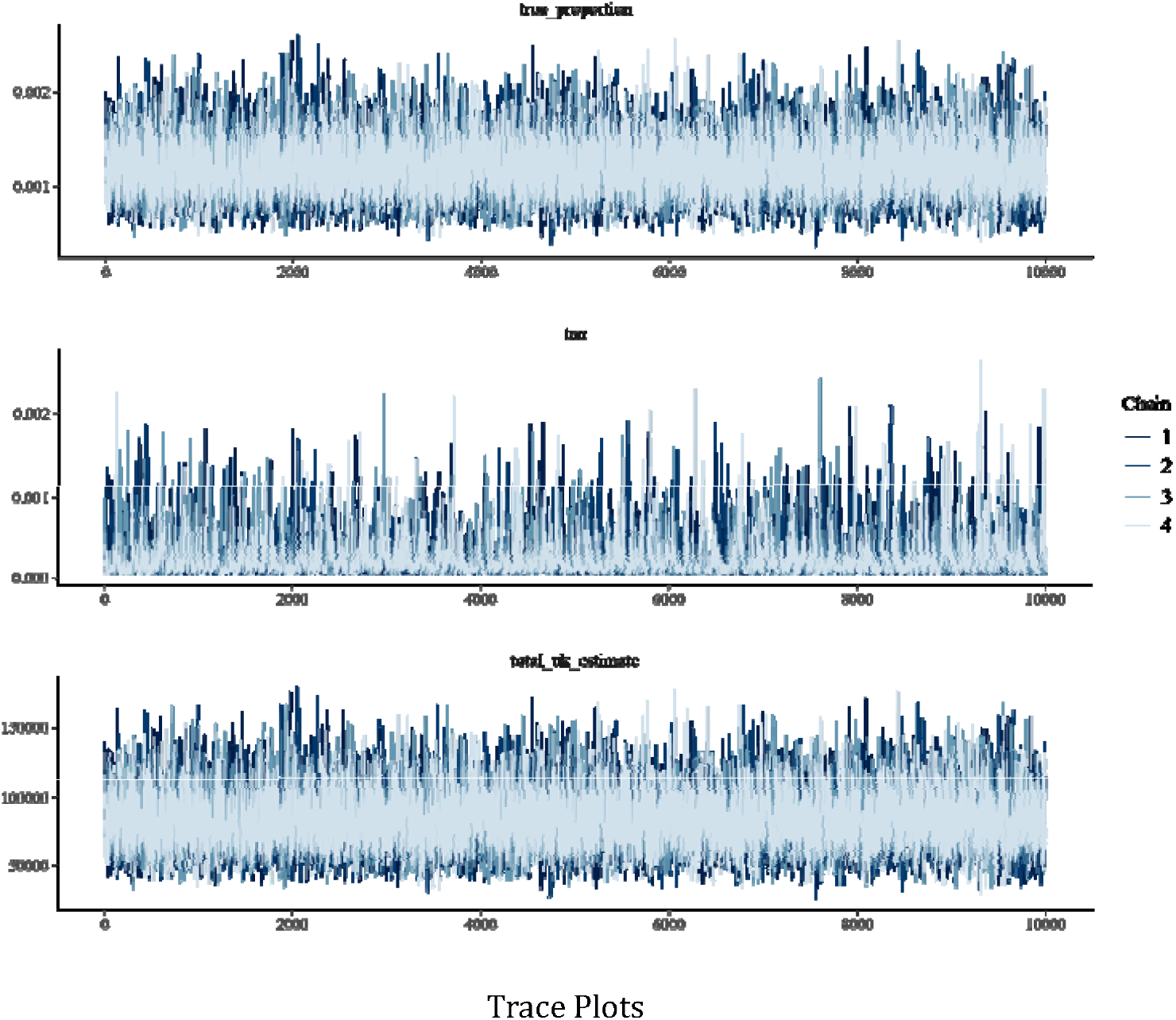

Critically, the model produced zero divergent transitions across 160,000 total sampling iterations (40,000 post-warmup). Divergent transitions indicate numerical instability and can compromise posterior inference; their absence demonstrates that the model successfully navigated the complex, high-dimensional parameter space without computational difficulties.

## Notes

### Competing Interest Statement

The authors have declared no competing interest.

### Author Declarations

The GUMCAD data was provided to us as aggregate figures, so it was already non-identifiable when we accessed it.

